# Prevalence of plantar ulcer and its risk factors in leprosy: a systematic review and meta-analysis

**DOI:** 10.1101/2023.08.17.23294254

**Authors:** Karthikeyan Govindasamy, Joydeepa Darlong, Samuel I Watson, Paramjit Gill

**Affiliations:** Warwick Centre for Global Health, Warwick Medical School, University of Warwick, Coventry, CV4 7AL, UK; Research Domain, The Leprosy Mission Trust India, New Delhi, India; Institute of Applied Health Research, College of Medical and Dental Sciences, University of Birmingham, Birmingham, B15 2TT, UK

**Keywords:** Plantar ulcers, leprosy, Prevalence of ulcer, plantar pressure

## Abstract

**Background:** Plantar ulcers are a leading complication of leprosy that requires frequent visits to hospital and is associated with stigma. The extent of burden of ulcers in leprosy and its risk factors are scant impeding the development of targeted interventions to prevent and promote healing of ulcers. The aim of this review is to generate evidence on the prevalence of plantar ulcer and its risk factors in leprosy.

**Methodology/Principal findings:** Databases (Medline, Embase, Web of Science, CINAHL, BVS), conference abstracts and reference lists were searched for eligible studies. Studies were included that reported a point prevalence of plantar ulcer and/or its “risk factors” associated with development of ulcers (either causatively or predictively), including individual level, disease related and bio-mechanical factors. We followed PRISMA guidelines for this review. Random-effects meta-analysis was undertaken to estimate the pooled point prevalence of ulcers. Reported risk factors in included studies were narratively synthesised. This review is registered in PROSPERO: CRD42022316726.

Overall, 15 studies (8 for prevalence of ulcer and 7 for risk factors) met the inclusion criteria. The pooled point prevalence of ulcer was 34% (95% CIs: 21%, 46%) and 7% (95% CIs: 4%, 11%) among those with foot anaesthesia and among all people affected by leprosy, respectively. Risk factors for developing ulcers included: unable to feel 10 grams of monofilament on sensory testing, pronated/hyper-pronated foot, foot with peak plantar pressure, foot with severe deformities, and those with lower education and the unemployed.

**Conclusions/Significance:** The prevalence of plantar ulceration in leprosy is as high as 34% among those with loss of sensation in the feet. However, the incidence and recurrence rates of ulceration are least reported. The inability to feel 10 grams of monofilament appears to be a strong predictor of those at risk of developing ulcers. However, there is a paucity of evidence on identifying those at risk of developing plantar ulcers in leprosy. Prospective studies are needed to estimate the incidence of ulcers. Identifying individuals at risk of ulcers will help design targeted interventions to minimize risk factors, prevent ulcers and promote ulcer healing.

## Introduction

Leprosy is a neglected and often stigmatised disease. It is more prevalent among those of young and middle age [1] and affected individuals may have to live with life-long disability due to the chronic nature of its impairments. Annually, 30% to 40% of newly diagnosed leprosy patients develop disabilities.[2] It is estimated that the prevalence of grade 2 disability according to WHO classification of impairments (visible impairments at the time of diagnosis) due to leprosy to be over 1 million worldwide by the year 2020.[3] The Posterior Tibial nerve damage has been reported to be affected in as high as 68% in people affected by leprosy [4] with varying rates among new cases [5-7] leading to loss of sensation in the foot and development of plantar ulcers. The recurrence of such ulcers leads to destruction of the bones of the foot, subsequent amputation [8], and in some squamous cell carcinoma [9]. The presence of ulcer is also associated with stigma [10] and the recurrent nature of ulcers affects participation in social life of the affected person [11]. While plantar ulcers can be prevented with appropriate and timely intervention [12-14], the lack of epidemiological literature describing the incidence, prevalence and factors associated with the plantar ulcer, limits the scope for designing and evaluating preventative interventions[15]. Reliable data on the prevalence and incidence of plantar ulcer will serve as baseline to study the effects of the interventions and compare between the interventions.

The aetiology of plantar ulcer is multifactorial. The loss of protective sensation in particular (defined as an inability to feel 10 grams of force),[16] foot deformities, and a resulting abnormal pressure distribution in plantar aspect of the foot are some of documented risk factors for ulcer [17, 18]. The role of occupation, activity level and other demographic factors are not known. Better understanding of the prevalence and risk factors for neuropathic ulcer development would help identify those at risk to provide targeted interventions, such as the provision of foot orthoses [19], customized footwear modifications, and self-care interventions at the community level. Therefore, we conducted a systematic review of primary epidemiological studies in people affected by leprosy with the aim of answering the questions: 1) What is the prevalence of plantar ulcer in the foot among people affected by leprosy and 2) What are the risk factors for development of plantar ulcers among people affected by leprosy?

## Methods

### Protocol and registrations

This review was registered on PROSPERO (CRD42022316726) and the protocol is provided in the supplementary information (S1 Text). We adapted the PRISMA guideline [20] for this review [21].

### Search strategy and study selection

We performed searches in the following databases MEDLINE, EMBASE, Web of science, CINAHL, and Biblioteca Virtual de Saude. In addition, we looked for unpublished resources like conference abstracts and screened reference lists of identified studies to be included in the review. We contacted the corresponding author of the selected studies to obtain full texts or for obtaining additional data. We used three keywords: ‘leprosy’, ‘ulcers’ and ‘risk factors’, along with related terms, in the search strategies combining medical subject headings (MeSH) and keywords. The search strategy is shown in the Supplementary information (S2 Text).

We included studies of any design that reported a point prevalence of plantar ulcer among people affected by leprosy. There was no restriction based on geographical location of the studies being carried out. We included studies from 1990 as a focus on disability prevention gained importance during this period after the introduction and success of multi-drug therapy in controlling the disease in 1983. Studies were excluded if they were case reports, editorials, letter to editors, qualitative studies and without available full texts. Articles in foreign languages where an English translation was not available were excluded.

The results of all searches were saved using reference manager software (Endnote 20) to identify and remove duplicates. Remaining articles were screened for inclusion in the review by screening title and abstract to be included for full text review. Two authors (KG and JD) screened studies and extracted data independently. A third reviewer checked and resolved discrepancies and disagreements.

### Quality assessment

The quality of included studies was assessed using the risk of bias tools for prevalence studies recommended by Hoy, Brooks [22]. To assess quality for studies of nonrandomized study designs such as before-after studies, cohort studies and case-control studies (only for risk factor part of the review), we used Risk of Bias Assessment tool for Nonrandomized Studies (RoBANS) [23]. The quality assessment was done independently by two authors (KG and JD) and discrepancies were resolved through consensus. All the eligible studies were included in the final analysis to calculate the pooled prevalence of plantar ulcer. We did not exclude any studies based on assessment of risk of bias but were used to categorize eligible studies as low to moderate risk and high risk to perform a sub-group analysis.

### Data extraction

Data were extracted into standardized database in MS Excel spreadsheet. For the prevalence of plantar ulcer, the data extraction variable included author name, year of publication, region/country (the area where the study was conducted), study setting (hospital/community), rural area/urban area, study design, sample size, study population, number of participants with loss of sensation in the foot and number of participants with outcome (ulcer). For the risk factors for plantar ulcers, we extracted relevant data for narrative synthesis using quantitative data on risk factor for ulcer development which was categorized as causative and predictive variables as shown in the Table 1.

**Table 1:** Risk factors for neuropathic ulcer in leprosy.

| Variables | Causative | Predictive |
| --- | --- | --- |
| Demographic |  | • Higher age group<br>• Gender<br>• Occupation<br>• Education level |
| Disease related and bio-mechanical factors | • Loss of touch sensation in the sole of the foot<br>• Higher vibration perception threshold<br>• Loss of sweating | • Previous ulcer<br>• High plantar pressure<br>• Foot drop<br>• Gross damage to bones of the joint<br>• callus/cracks<br>• non-adherence to wearing footwear / self-care<br>• Duration of disease |

### Data synthesis and analysis plan

The point prevalence of plantar ulcer in the feet was calculated using number of persons reported in the study with plantar ulcer as numerator. We reported the point prevalence of neuropathic plantar ulcer with two different denominators as 1) among people with loss of sensation in the foot due to leprosy and 2) among all people affected by leprosy. Where only a prevalence is reported and not the numerator or denominator data, we looked for the definition of the denominator population to ascertain. We calculated the 95% confidence intervals using the standard binomial formula. We conducted a random-effects Beta-binomial meta-analysis to estimate the pooled point prevalence of neuropathic ulcer in the two populations (in those with neuropathy and in all patients affected by leprosy). We estimated the *I^2^* statistic to examine between-study heterogeneity. We presented the pooled analysis as forest plot for all eligible studies and disaggregated based on the risk of bias assessment as low to moderate risk and high risk of bias.

We conducted a narrative synthesis of quantitative data on risk factors for development of neuropathic plantar ulcer. We anticipated heterogeneity between studies in their methodological, statistical, and clinical approaches. For example, the plantar pressure was reported as continuous variable as peak plantar pressure or as binary/ordinal variables as low, normal, and high as compared to normal foot or foot with loss of sensation but without ulcer. Similarly, the loss of sensation in the foot may be ascertained using monofilament or ball-point pen. These risk factors were examined in heterogeneous patient groups. We stratified and combined the studies based on risk factors, population, and method of analysis. We then narrated the association between these factors with the outcome (ulcer) to synthesise evidence.

## Results

Our search returned a total of 1,397 studies; of them 1,345 were excluded after removal of duplicates (505) and screening of title and abstract (830), leaving 62 eligible full text articles. We excluded 50 studies (S3 Text), leaving eight articles reporting prevalence of ulcer and seven articles reporting risk factors for ulcer were included in the final review. A detail of eligible studies included in the review are shown in the PRISMA flow chart (Fig 1). The summary of included studies to estimate the prevalence of ulcer is shown in Table 2. All studies included in the review of prevalence of plantar ulcer were hospital-based studies except one from community-based setting.

**Figure 1:**
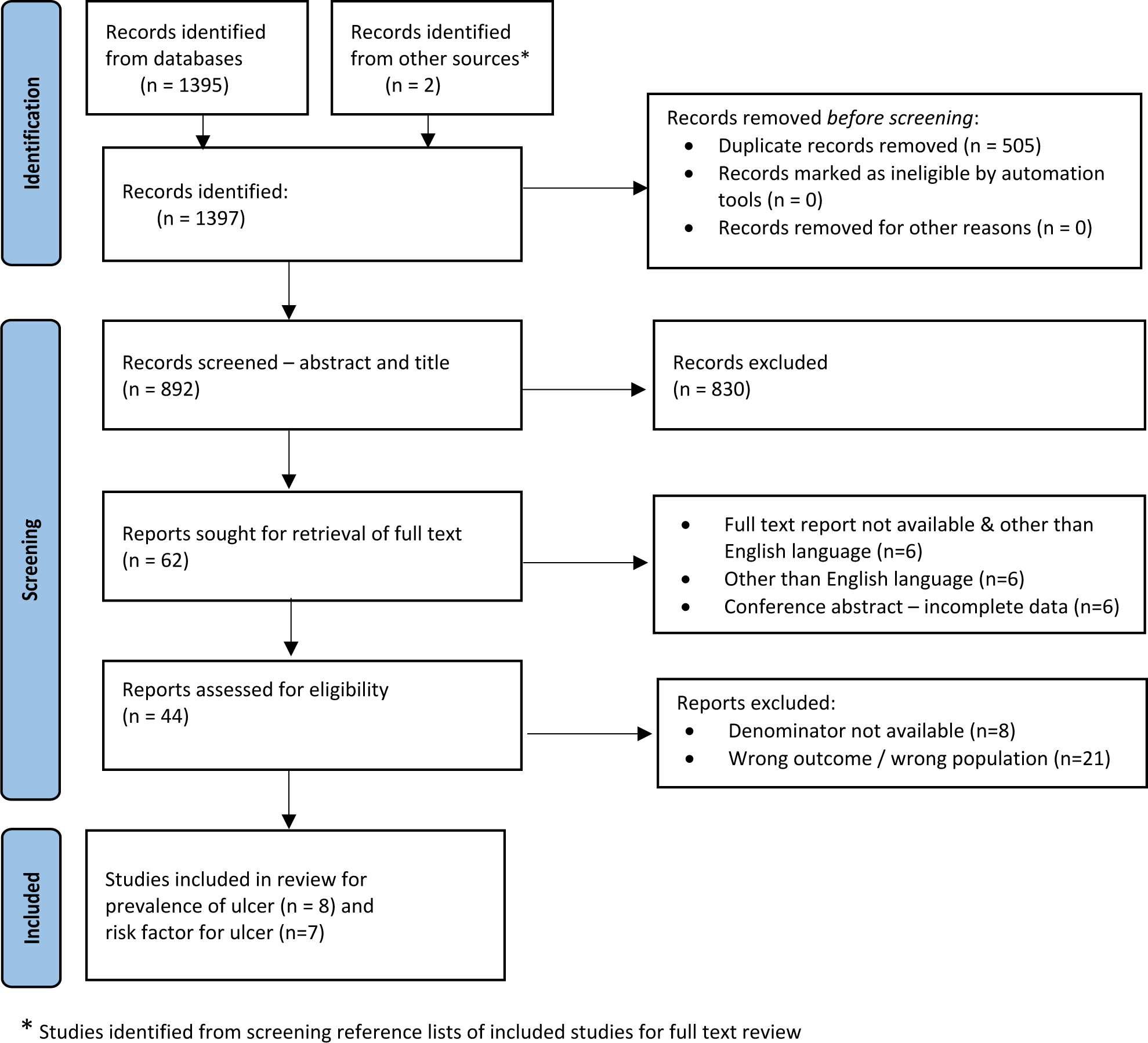
Preferred Reporting Items for Systematic Reviews and Meta-Analyses (PRISMA) flow diagram of the study selection process.

**Table 2:**
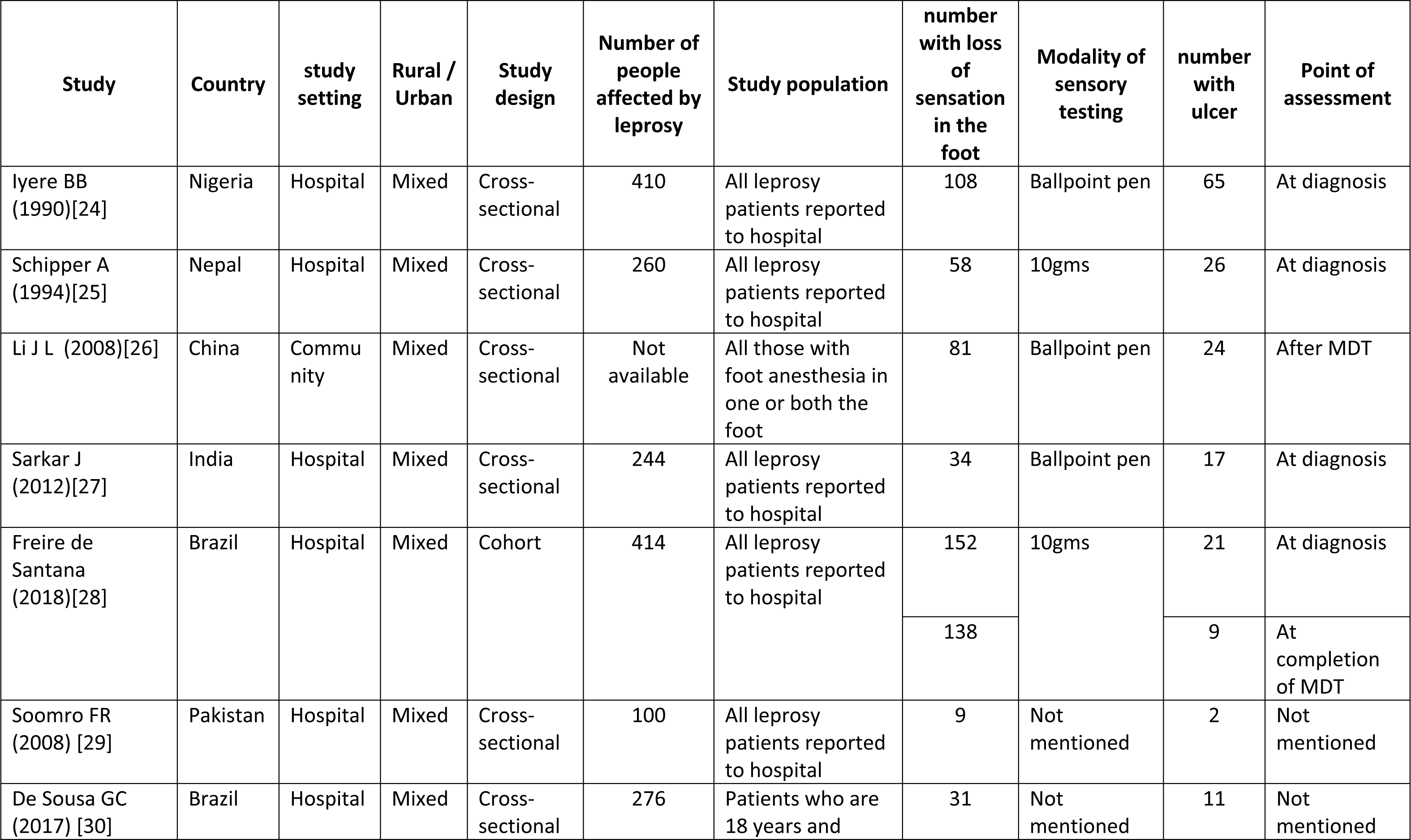
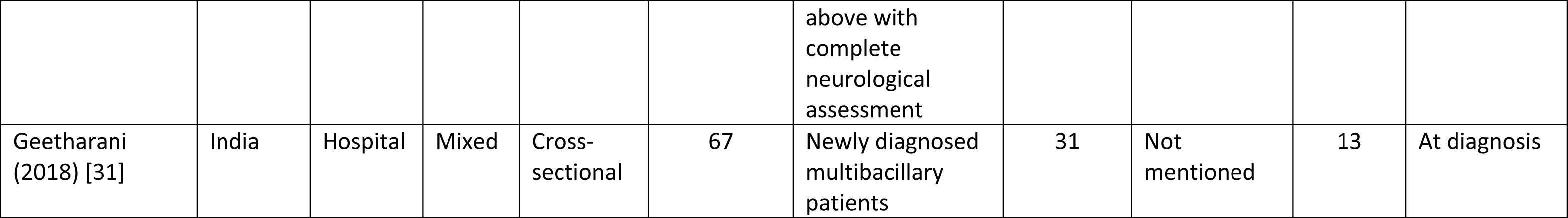
Summary of studies included in the review for prevalence of ulcer.

### Prevalence of ulcer

Eight studies reported a point prevalence of plantar ulcer among those at risk (with loss of sensation in the foot). Four out of eight studies included prevalence data collected at the time of diagnosis of leprosy,[24],[25],[27],[31] one study at both diagnosis and completion of medical treatment for leprosy[28] and in one study data was collected post completion of medical treatment.[26] While, in two studies point of data collection was not mentioned and they included all patients reported to hospital during the study period[29],[30]. The estimated pooled point prevalence of ulcer among those at risk was 34% (95% CIs 21%, 46%) shown in Fig 2. There was high heterogeneity (I^2^= 95%, p-value= <0.01). In the sub-group analysis based on the risk of bias assessment, the pooled prevalence from studies with low to moderate risk of bias was 34% (95% CIs 16%, 51%) while those studies with high risk of bias was 36% (95% CIs, 25%, 47%)

**Figure 2:**
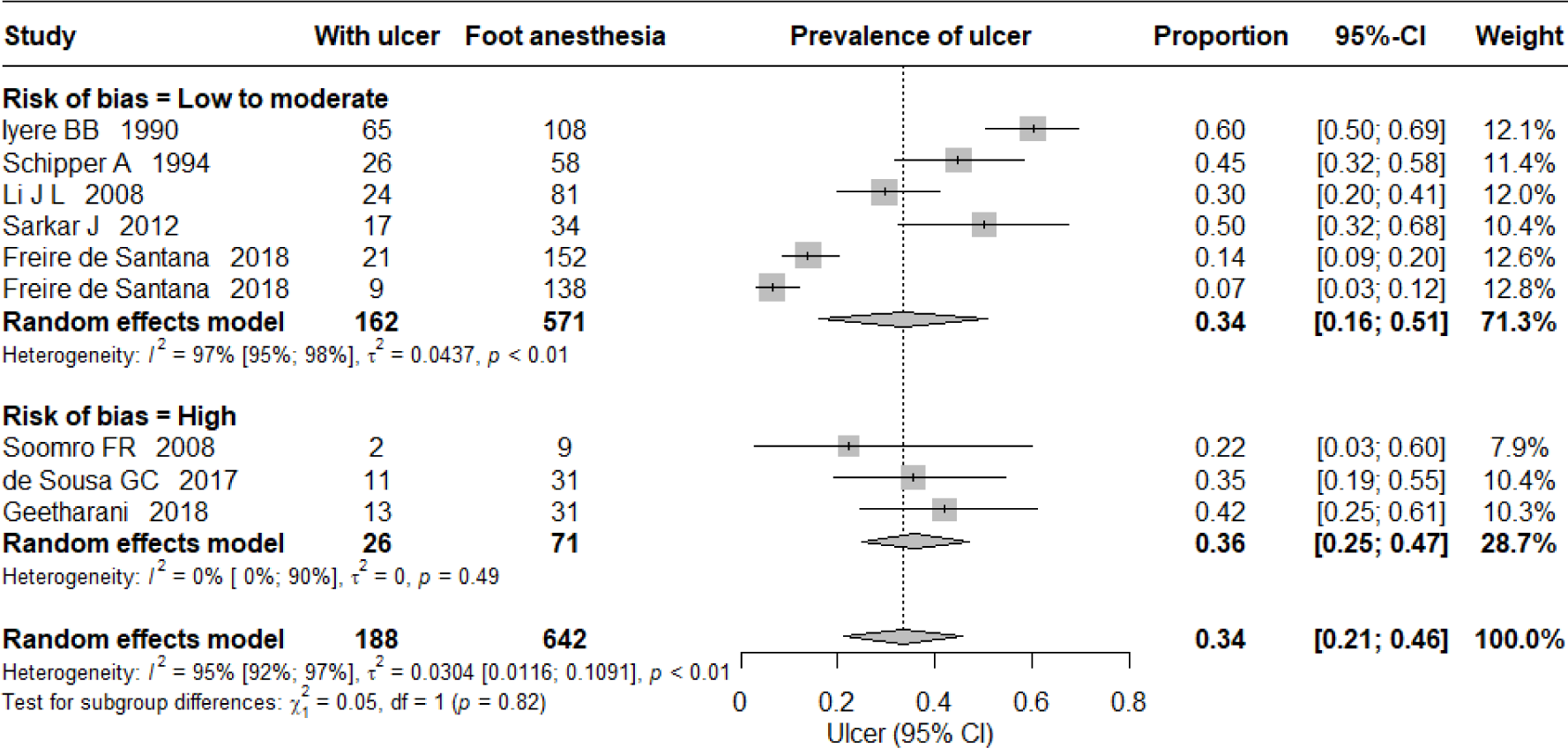
Prevalence of ulcer among those at risk (with loss of sensation in the foot)

To estimate the point prevalence of ulcer among all people affected by leprosy Li J L et al (2008) study was excluded as their study population comprised only those at risk (with loss of sensation in the foot) identified from the community settings. The estimated pooled point prevalence of ulcer among all people affected by leprosy was 7% (95% CIs 4%, 11%) and it is shown in the Fig 3. There was a high heterogeneity I^2^= 91%, p-value = <0.01). In the sub-group analysis based on the risk of bias assessment, the pooled prevalence from studies with low to moderate risk of bias was 8% (95% CIs 3%, 12%) while those studies with high risk of bias was 8% (95% CIs, 0%, 17%).

**Figure 3:**
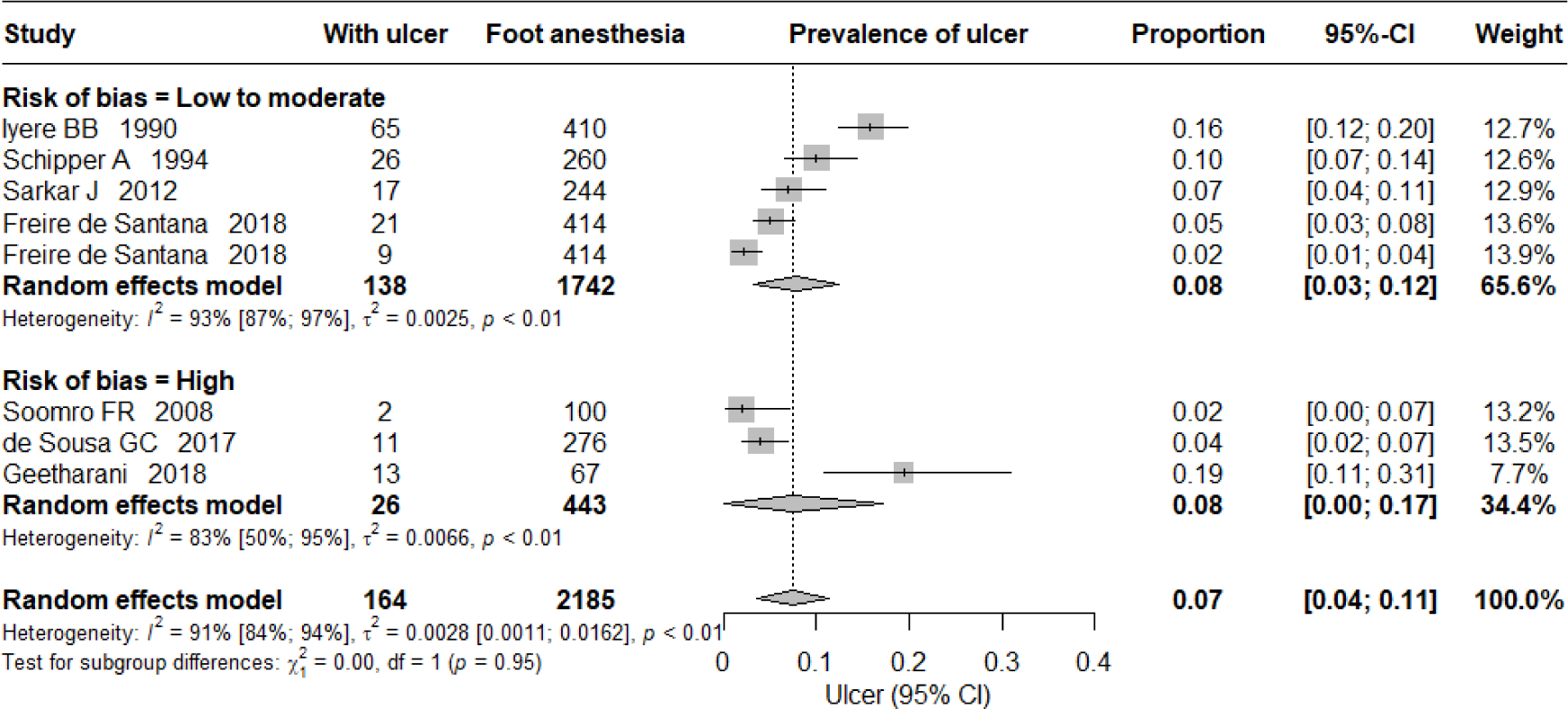
Prevalence of ulcer among all people affected by leprosy.

### Risk factors for ulcer

Seven studies were included in the final review of risk factors for development of foot ulcers. Five studies were cross-sectional study design and two were case-control studies. Two studies assessed demographic and clinical factors,[32],[33] three studies assessed clinical factors[34],[35],[36] and two studies assessed the bio-mechanical factors.[37],[38]. The summary of included studies is given in Table 3.

**Table 3.** Summary of studies included for risk factors.

| Study | Country | study setting | Rural / Urban | Study design | Sample size | Study population | number with loss of sensation in the foot | Modality of sensory testing | number with ulcer | Point of assessment | Risk factors assessed |
| --- | --- | --- | --- | --- | --- | --- | --- | --- | --- | --- | --- |
| Mustapha G (2019)[32] | Nigeria | Hospital | Mixed | case-control | 124 patients with ulcer and 118 without ulcer | 242 patients with leprosy | 233 patients | Ballpoint pen | 124/242 patients | Mixed | Treatment status, age, gender, occupation, educational status, utilization of footwear, ulcer relapse and knowledge on self-care |
| Kunst H (2000)[33] | Pakistan | Hospital | Mixed | cross-sectional | 55 patients with ulcers | Patients admitted in hospital for ulcer care | 55 patients | Wisp of cotton | 40/55 patients | Post MDT | Age, gender, duration of leprosy, physical deformity level |
| Malaviya GN (1997)[34] | India | Hospital | Mixed | cross-sectional | 40 patients (80 feet) | People affected by leprosy with loss of sensation in the hands and foot | 61 feet | Graded nylon thread | 22/240 sites in the foot | Mixed | Touch sensation - monofilament |
| Birke (2000)[35] | Thailand | Hospital | Mixed | cross-sectional | 101 patients, 61 with history of foot ulceration | People affected by leprosy with loss of sensation in the hands and foot | 101 patients | Graded nylon thread | Not reported | Mixed | Touch sensation- Monofilament |
| Feenstra W (2001)[36] | Ethiopia | Hospital | Mixed | case-control | 75 patients (43 with history or with ulcer and 32 never had ulcer) | People affected by leprosy with loss of sensation in the foot | 75 patients | 10 grams monofilament | 43/75 with history or with ulcer | Post MDT | Touch sensation, vibration sensation, Ankle ROM, Sub-talar joint position, Great toe extension and walking pattern |
| Cross H<br>(2007)[37] | Nepal<br>and<br>Philippines | Hospital | Mixed | cross-<br>sectiona<br>l | 144 subjects<br>(69 normal<br>foot, 75 feet<br>with loss of<br>sensation with<br>or without<br>ulcer) | People affected<br>by leprosy with<br>loss of sensation<br>in the foot | 75 feet with loss<br>of sensation<br>with or without<br>ulcer (263 foot<br>included in the<br>analysis) | 10 grams<br>monofila<br>ment | 39/110<br>feet | Mixe<br>d | Plantar pressure and<br>Subtalar joint position |
| Schie van<br>(2013)[38] | Netherl<br>ands | Hospital | Urban | cross-<br>sectiona<br>l | 15 with no<br>history of<br>ulcer, 15 with<br>previous ulcer,<br>9 with current<br>ulcer | People affected<br>by leprosy with<br>loss of sensation<br>in the foot | All 39 patients<br>included in the<br>study | Graded<br>nylon<br>thread | 9<br>patients<br>with<br>ulcer | Post<br>MDT | Barefoot peak pressure,<br>In-shoe peak pressure,<br>daily cumulative<br>forefoot stress, In-shoe<br>forefoot pressure time<br>integral, Daily strides |

#### Demographic factors

The occurrence of ulcer was higher among males, those with advanced age, lack of formal education [29] and unemployed [36].

#### Clinical factors

#### Sensory neuropathy

The loss of touch sensation detected by monofilament (inability to feel 7 grams or more) was associated with risk of plantar ulcers. In a study from Thailand reported that inability to feel 7 grams (filament number 4.19) of monofilament had 97% sensitivity and 100% specificity in identifying foot with ulcerations. The sensitivity and specificity increased to 100% when person was inability to feel 10 grams of force.[39] Similarly, a study from India found that none who could feel monofilament of 7 gram force or below had an ulcer. The risk of ulcer increases when the threshold of touch sensation was twice the normal threshold [34]. In a study from Ethiopia noted that about 43% (48/112) of points (part of the foot) where patient was unable to feel 10 grams of force had had ulcer or with ulcer which is lower than those other reports[36].

One study reported the associated risk when one is not able to feel the vibration sensation. Among different points in foot had had ulcer or with ulcer, only 47% (48/103) points had impaired vibration perception (256Hz)[36].

#### Foot deformities

Presence of any foot deformities increases the risk of ulcer. The foot deformities and its severity such as claw toes, footdrop, reduced medial arch and bone absorption increased a risk of ulcer as compared to those with only loss of sensation. The severity of foot deformities in turn was associated with the increased age and the duration of the disease.[33]

#### Bio-mechanical factors

Deviation from the neutral sub-talar joint position was strongly associated with the risk of ulcer. Pronated or hyper-pronated (subtalar joint) increased the risk of ulcer followed by supinated foot as compared to neutral subtalar joint as observed in a standing position. As a result, there was a corresponding increase in the plantar pressure which was associated with the increased risk of ulcer.[37]

The increase in the peak plantar pressure over vulnerable sites increased the risk of ulcer. It was observed that the barefoot peak plantar pressure was higher for those with ulcer or history of ulcer as compared to those who never had an ulcer. The in-shoe peak plantar pressure was considerably higher for those with ulcer as compared to those without ulcer. The daily cumulative stress over foot was not associated with the risk of ulceration as much as increased peak plantar pressure.[38]

## Discussion

### Prevalence of plantar ulcer

The burden of plantar ulcer in leprosy is always considered high but to the best of our knowledge its magnitude has not been reported and systematically studied. This systematic review summarised the current literature on the prevalence of plantar ulcer. From the eight studies included in the review the pooled prevalence of plantar ulcer among those at risk (loss of sensation in the foot) was 34% and among all people affected by leprosy it was 7%. As we expected, there was high variation between the studies included in the review. In the sub-group analysis based on risk bias, the pooled prevalence almost remained same (Fig 2 and 3). However, the confidence interval for the prevalence estimated among those at risk of ulcer was wider in studies with low to moderate risk of bias as compared to studies with high risk of bias. Whereas, in the pooled prevalence of ulcer among all those affected by leprosy, the confidence interval was wider for those studies with high risk of bias as compared to studies with low to moderate risk of bas. The main reason for excluding studies from the review was the lack of reporting of the size of the denominator population, so we could not calculate the prevalence.

The estimated pooled prevalence of plantar ulcer could perhaps be slightly higher than actual burden in the population as seven out of eight studies included were based in tertiary hospital for leprosy where people with advanced diseases are treated. In one study from Brazil, the prevalence was higher at the time of diagnosis (13%) than at the time of completion of Multi-drug therapy (MDT) within the same cohort (6%), likely because the patients were under supervision while on MDT.[28] However, the prevalence of ulcer was 29% when the study sample was drawn from the community among those completed MDT which may reflect the true prevalence in the population[26]. There is a decreasing trend in the prevalence of ulcer over a time based on the included studies. However, the decreasing trend observed may not reflect the actual burden of patients with ulcers in the community which is not known. It is likely that the burden could be considerably higher due to cumulative number of patients over the decades with loss of sensation in the feet would increase the number of patients with ulcer, Also, the recurrent nature of the foot ulcer will further add to the burden. None of the included studies reported incidence of the ulcer or severity of the ulcer in terms of its size. Nevertheless, the estimated prevalence may serve as baseline for studying the effectiveness of the self-care program aimed at reducing the prevalence of the ulcer. Data on severity of ulcer would be an essential component of the evaluation of self-care intervention as it will not only reduce the prevalence of ulcer but also promote healing of ulcer where change in its size will serve as an outcome.[40]

### Risk factors for developing plantar ulcer

Understanding the factors associated with plantar ulcers in leprosy is an essential step towards identifying those at high risk of ulcers for targeted preventive and curative intervention. The earliest discussions of the development of plantar ulcer emphasized that: 1) ulcers are not random and each ulcer has its own patho-mechanics, 2) ulcers start as blisters over areas of necrosis and 3) the ulcers develop because of infection through penetrating wound or infection of deep cracks and because of the stresses over the plantar tissues while walking leading to ulcers.[41-43] The first point demands the better understanding of those at risk of ulcer for the effective intervention. The current literature categorizes risk factors as pre-disposing factors (sensory loss, loss of sweating and foot deformities), pre-ulcerative conditions (corns, callus, fissures, blisters and haematoma) and direct causes (trauma, high shearing stress and high plantar pressure) that lead to plantar ulcers. [44, 45] But this categorization of risk factors does not help identify those at risk of ulcer and provide appropriate intervention to prevent from ulcer development and to promote healing when one develops ulcer. For example, when patient present with loss of sensation (inability to feel 10 grams of force) and pronated foot and increased plantar pressure over medial three meta-tarsal heads, they are likely to develop ulcer over pressure area. For such foot provision of medial arch support would help offload a pressure area and thus prevent from ulcer development. As a result, patients are rarely seen at the early stage of the ulcers in the routine practice. In the past unsuccessfully attempts has been made to determine those at risk of developing plantar.[34-36] In this review we attempted to identify those factor or combination factors that can identify those at risk of developing ulcers.

Among demographic factors, advanced age appears to be increasing the risk of plantar ulcers. The possible explanation could be that in the advanced age there is an alteration in the foot structure and function with the foot becoming flat and pronated. When coupled with loss of sensation, this could lead to excessive pressure in the forefoot resulting in ulcer.[46] Increased age also was associated with a longer duration of disease / impairments, which could further increase the risk of plantar ulcer.[32]. This observation is consistent with evidence on diabetic foot ulcers, where the occurrence of ulcer was higher among those 50 years and above.[47, 48]

Men with leprosy disproportionately develop plantar ulcers.[32] The incidence of leprosy is higher among males than females[49], hence the complications including ulcer could be higher among male. The male predisposition is also observed in diabetes foot ulcer.[47, 48, 50, 51] Possibly, men do more walking and rigorous physical activity for their social role as earning member as compared to female.[52] Mustapha G et al reported that the unemployment and poor socio-economic status was associated with developing ulcers but their role is not clear.[32] However, their study was the case-control design limiting the inference of causality and it is possible that leprosy and its complications like ulcer can cause stigma and limit ability to work which can lead to poor socio-economic status therefore, here the direction of causality is unclear. Prospective investigation could better explain the role of these factors.

Among clinical factors, inability to feel monofilament of 7 grams force is found to be a most sensitive (97%) and specific (100%) predictor of ulcer development, the sensitivity increases to 100% when one is unable to feel 10 grams of force. Inability to feel 10 grams of force is considered loss of protective sensation, hence increasing the risk of ulcer. [53] In Feenstra study, only 43% of points (sub-part of the foot) in the foot had impaired sensation according to 10 grams which is too low as compared to other studies.[36] This may be due to taking sub-parts of the foot than whole foot to determine the loss of sensation. It is important to note here that high sensitivity of 7 grams may indicate that many with some amounts of intact protective sensation do develop ulcers and by the time patient’s ability to feel 10 grams could be too late to prevent ulcers. It is possible that patients may have protective sensation but impaired temperature sensation; unable to feel warm and cold, which could also lead to ulcer. Impaired warm sensation precedes in the sub-section of the people affected by leprosy even before the (touch) sensory loss is clinically evident when tested using monofilaments.[54] Nonetheless, the validity of using a monofilament for sensory testing has been established well in a large cohort study.[7] Leprosy programs still use a ballpoint pen to test the sensation instead of monofilaments in routine leprosy clinics which could be too late to prevent ulceration.[55] The loss of vibration sensation was found to be associated with increased risk for ulcer.[36] The method of determining the vibration perception impairment was qualitative where person is asked if he/she can feel the vibration given using tuning fork, where the actual threshold to perceive the vibration may be lower than the given vibration frequency. Also, in this study, multiple parts of the foot than the whole foot was included in the analysis, which could be a reason for low sensitivity in identifying foot at risk for ulcer.

Among seven studies included in the review two studies reported biomechanical factors where the pronated or hyper-pronated foot were associated with increased plantar pressure which in turn was associated with ulcer.[37, 38] van Schie et al reported that higher barefoot and in-shoe plantar pressure were associated with the presence of ulcer or history of ulcer. The evidence suggest that increased peak plantar pressures lead to tissue breakdown overtime causing ulcer in both leprosy and diabetic foot ulcers.[56, 57] While cumulative activity level was not associated with the risk of ulcer, indicating that the risk is likely to be a combination of foot structure and function than the activity level. Plantar pressure was measured in the included studies in static (standing) position which may be different from pattern observed while walking. Given the advancement in the technology it is possible to assess during dynamic (while walking) position which will be more accurate than the static position. The simple podiatry assessments [58] can identify foot alignment which can indicate abnormal plantar pressure distribution in the foot.[59] The podiatry assessment can help not only to identify person at risk of ulcer but also help provide podiatric intervention which will reduce the impact of high plantar pressures. [19, 60, 61] The podiatry interventions are found to be effective in reducing the plantar ulcers in diabetes[62] but are least utilised in leprosy. The prospective studies are needed to determine the role of biomechanical factors in identifying person at risk of ulcer for a focused podiatry intervention.

History of previous ulceration as risk of future ulcers is not reported in leprosy, but found to be a strong predictor of ulcer in diabetes.[63] None reported the duration of disease, lifestyle, body mass index which are likely to contribute along with neuropathy, foot structure and function.

It is important note from the included studies that each study looked at specific categories of risk factors, either demographic and/or clinical variables or bio-mechanical factors. The risk of ulcer is likely to be combination various factors that needs to be investigated in a prospective study.

### Strength and limitations

This review adopted a thorough search strategy. To improve the rigour of the review two independent reviewers were involved in the process of screening of articles, risk of bias assessment and data extraction. A few limitations include that many included studies were from referral hospitals which may over represent those with severe form of disease, therefore, the estimated prevalence of ulcer may be higher than the population value.

There was a high heterogeneity as included studies varied in their sample size and study populations included from different geographic locations. The number of studies included for review of risk factor for ulcer are insufficient to calculate risk ratios. Also, studies are either cross-sectional or case-control study which limits the causal inferences of the identified risk factors. We could not study the role of social and lifestyle related determinants for ulcer development which are equally as important as clinical and biomechanical factors. There is a need for a population-based survey to estimate the prevalence of ulcer. Prospective studies on risk factors would help better understand the causation of ulcer and develop targeted interventions.

## Conclusions

The prevalence of plantar ulceration in leprosy is as high as 34% among those with loss of sensation in the feet. However, the incidence and recurrence rates of ulceration are least reported. The inability to feel 10 grams of monofilament appears to be a strong predictor of those at risk of developing ulcers. However, there is a paucity of evidence on identifying those at risk of developing plantar ulcers in leprosy. Prospective studies are needed to estimate the incidence and prevalence of ulcers. Identifying individuals at risk of ulcers will help design targeted interventions to minimize risk factors, prevent ulcers and promote ulcer healing.

## Supporting information

S1 Text - PROSPERO registration

S2 Text – Search strategy

S3 Text – List of excluded studies

## Data Availability

All relevant data are within the manuscript and its Supporting Information files.

## Authors summary

Foot ulcers are a common complication for individuals with loss of foot sensation due to leprosy and are a primary source of stigma. This review found that approximately 34% of those suffering from such sensory loss, and 7% of all those affected by leprosy, have ulcers. These figures can provide a baseline for evaluating ulcer prevention or healing interventions. However, lack of data on the incidence and recurrence of ulcers impedes a comprehensive understanding of the burden of foot ulcer. The underlying process for plantar ulcer development in leprosy remains least studied, restricting the potential for targeted preventative measures. This review noted that males and those of lower socio-economic status afflicted with leprosy are more prone to ulcer development. An inability to perceive a force of 10 grams (touch sensation) on the sole of the foot found to be a sensitive indicator for ulcer development. Inward rolling of the foot while walking, which increases pressure on the foot’s inner side, also heightens ulcer risks on these high-pressure areas. Future studies will illuminate the ulcer development process, aiding in identifying individuals at higher risk of developing ulcers.

## Acknowledgments

We thank Samantha Johnson, academic support librarian, Warwick Medical School, University of Warwick for her support in developing the search strategy. This review was funded by the National Institute for Health and Care Research (NIHR) (NIHR200132) and from the UK Government to support global health research and PG is NIHR Senior investigator. The views expressed in this publication are those of the authors and not necessarily those of the NIHR or the UK Department of Health and Social Care.

